# Partial Verification Bias Correction Using Scaled Inverse Probability Resampling for Binary Diagnostic Tests

**DOI:** 10.1101/2025.03.09.25323631

**Authors:** Wan Nor Arifin, Umi Kalsom Yusof

## Abstract

Diagnostic accuracy studies are crucial for evaluating new tests before their clinical application. These tests are compared against their respective gold standard tests, and accuracy measures such as sensitivity (Sn) and specificity (Sp) are often calculated. However, these studies frequently suffer from partial verification bias (PVB) due to selective verification of patients. PVB eventually leads to biased accuracy estimates in such studies. Among methods developed for PVB correction under the missing at random assumption for binary diagnostic tests, a bootstrap-based method known as the inverse probability bootstrap (IPB) was proposed. Despite showing low bias for estimating Sn and Sp, the IPB method exhibited higher standard errors than other PVB correction methods. This paper introduces two new methods: scaled inverse probability weighted resampling (SIPW) and scaled inverse probability weighted balanced resampling (SIPW-B), which build upon the IPB approach. Through simulations and clinical data, SIPW and SIPW-B were compared against IPB and other methods. The results demonstrated that the new methods outperformed IPB by showing lower bias and standard errors in Sn and Sp estimation. Specifically, SIPW-B outperformed IPB in Sn estimation, while SIPW performed better in Sp estimation, particularly when disease prevalence is low. These methods offer advantages such as complete data restoration and calculations independent of disease prevalence. Although computationally demanding, this limitation becomes less significant with the increasing power of modern computing resources.

## Introduction

Diagnostic tests are crucial in medical care, so ensuring their clinical validity through diagnostic accuracy studies is essential [1, 2]. These studies involve comparing a new test with an established gold standard to evaluate its performance using accuracy measures like sensitivity (Sn) and specificity (Sp) for binary tests [1, 3–5]. However, verifying disease status with the gold standard can be expensive, time-consuming, and invasive [1, 5–8]. This verification challenge often leads to partial verification bias (PVB), where patients with positive test results are predominantly selected for gold standard verification, while fewer patients with negative test results are verified [1, 6, 8, 9]. This gives rise to a missing at random (MAR) missing data mechanism, as the decision to verify depends on the result of the diagnostic test [5, 6].

It is important to correct the bias during the analysis as PVB leads to biased accuracy measures in diagnostic tests [1, 6, 10]. PVB correction methods have been comprehensively reviewed by [2] and [11]. For binary tests under the MAR assumption, PVB correction methods can be categorized into Begg and Greene’s (BG)-based, propensity score (PS)-based, and multiple imputation (MI) methods. BG-based and MI methods adjust accuracy measures by calculating probability of disease status given test result, which are unbiased under MAR assumptions [3, 12]. In contrast, PS-based methods estimate the probability of verification given test result and use weighting to correct the bias [13, 14]. Several medical studies have applied these correction methods in their research, signifying the importance of the bias correction [15–19].

Inverse probability bootstrap (IPB) sampling was introduced to address sampling bias in model-based analyses [20]. While the bootstrap technique is traditionally used for estimating standard errors, IPB leverages this technique to achieve unbiased parameter estimates by creating weighted samples. This method corrects the sample distribution without requiring extensive modifications or new methods [20]. Because it is a bootstrap approach, IPB simplifies the estimation of standard errors and enables the calculation of confidence intervals for statistical inference [20].

The PS-based method for PVB correction and IPB share a common approach, where they both begin by estimating the selection probability, or verification probability in the context of PVB, and then use this probability to correct bias through weighting methods. The IPB approach offers an attractive way to address bias due to its reliance on the bootstrap technique, which comes with several advantages. Therefore, IPB was adapted in the context of PVB correction in a study by Arifin and Yusof [21]. In the study, IPB demonstrated low bias for estimation of Sn and Sp. However, it showed relatively higher SE than other PVB correction methods and it only corrects the distribution of the verified portion of the PVB data. This study proposed two methods namely scaled inverse probability weighted resampling (SIPW) and scaled inverse probability weighted balanced resampling (SIPW-B) to overcome these limitations with IPB in the context of PVB correction under the MAR assumption for binary diagnostic tests.

## Materials and methods

The simulated and clinical data sets used in this study, the proposed methods for PVB correction based on IPB, the metrics for performance evaluation, the selected methods for comparison, and the experimental setup of this study are described in this section. The followinf notations are used: *T* = test result, *D* = disease status, *V* = verification status, *n*_1_ = verified observations, *n*_0_ = unverified observations, and *n* = all observations, or *n*_1_ + *n*_0_.

### Data sets

This study evaluated and compared different methods using both simulated and clinical datasets. Simulated data allowed for performance assessment against known parameter values [20, 22], while clinical data enabled comparisons based on established reference data, mirroring the approach used in previous PVB correction research [23–26].

#### Simulated data sets

The simulated data sets were generated using the settings described in [21]. The settings were adapted from [24], [25] and [26]. The settings are described as follows:

1. True disease prevalence (*p*) or *P*(*D* = 1): moderate = 0.40 and low = 0.10.
2. True sensitivity (Sn) *P*(*T* = 1|*D* = 1): low = 0.3, moderate = 0.6, high = 0.9
3. True specificity (Sp) *P*(*T* = 0|*D* = 0): moderate = 0.6, high = 0.9.
4. Verification probabilities: When the verification depends only on test result, this is a MAR missingness mechanism. Fixed verification probabilities given test result *P*(*V* = 1|*T* = *t*) were set at *P*(*V* = 1|*T* = 1) = 0.8 and *P*(*V* = 1|*T* = 0) = 0.4 [24]. In words, when test results are positive, patients are more likely to be verified with probability of 0.8. On the other hand, when test results are negative, patients are less likely to be verified with probability of 0.4.
5. Sample sizes, *n*: 200 and 1000.

For the complete data, the probability of counts in the 2 *×* 2 cross-tabulated table of *T* versus *D* are distributed as a multinomial distribution [24, 26]. Based on pre-specified Sn = *P*(*T* = 1|*D* = 1), Sp = *P*(*T* = 0|*D* = 0) and *p* = *P*(*D* = 1) = *π*, the probabilities of counts are distributed as *M*(*π*_1_, *π*_2_, *π*_3_, *π*_4_), where

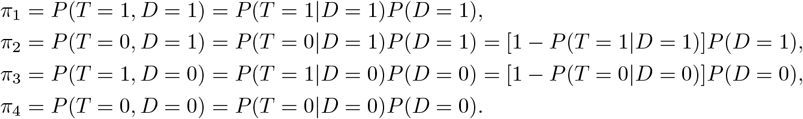

For each specified sample size *n*, generating a simulated data set with MAR-induced PVB involves the following steps:

1. A complete data set of size *n*, distributed as multinomial distribution, *M*(*π*_1_, *π*_2_, *π*_3_, *π*_4_) was generated. Numerical value were randomly generated from 1, 2, 3, 4 according to these probabilities.
2. The values were converted into realizations of *T* = *t* and *D* = *d* variables, where the numbers were mapped as: 1 *→*(*T* = 1, *D* = 1), 2 *→*(*T* = 0, *D* = 1), 3 *→*(*T* = 1, *D* = 0) and 4 *→*(*T* = 0, *D* = 0).
3. Under the MAR assumption, the PVB data set was generated by adding *V* = {1, 0} with verification probability of *P*(*V* = 1|*T* = 1) = 0.8 and *P*(*V* = 1| *T* = 0) = 0.4, where *V* follows binomial distribution. *D* values for *V* = 0 observations were set as *NA* to create missing values.

The simulated data layout for the full data and PVB data sets is shown in Figure 1.

**Fig 1.**
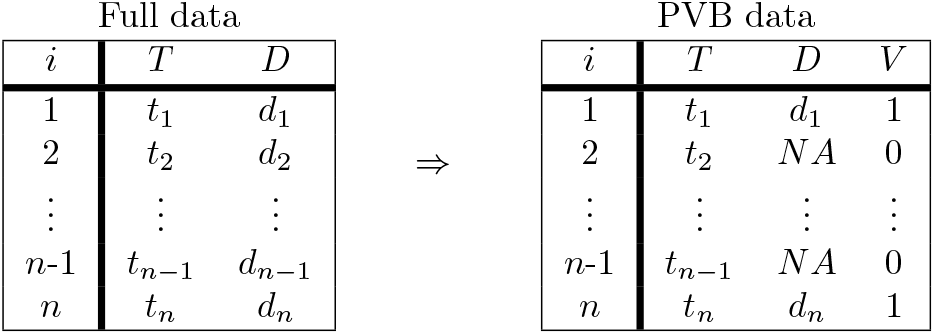
Simulated data layout for full and PVB data.

#### Clinical data sets

This study utilized two commonly used clinical data sets from [24, 26–30] to show and compare the implementation of the PVB correction methods in real-world situations. The original data obtained from the cited studies were converted to an analysis-ready format (.csv). These data sets are described as follows:

##### 1. Hepatic scintigraphy test

The data set pertains to hepatic scintigraphy, a diagnostic imaging technique used for detecting liver cancer, as provided in the original study [27]. The test was performed on 650 patients, where 344 patients were verified by liver pathological examination (gold standard test). The percentage of unverified patients is 47.1%. The data set contains the following variables:

a. **Liver cancer**, *disease*: Binary, 1 = Yes, 0 = No
b. **Hepatic scintigraphy**, *test*: Binary, 1 = Positive, 0 = Negative
c. **Verified**, *verified*: Binary, 1 = Yes, 0 = No

##### 2. Diaphanography test

The data set pertains to diaphanography test for detection of breast cancer, as provided in the original study [28]. Diaphanography test is a noninvasive method (diagnostic test) of breast examination by transillumination using visible or infrared light to detect the presence of breast cancer. The test was performed on 900 patients. Only 88 patients were verified by breast tissue biopsy for histological examination (gold standard test). The percentage of unverified patients is 90.2%. The data set contains the following variables:

a. **Breast cancer**, *disease*: Binary, 1 = Yes, 0 = No
b. **Diaphanography**, *test*: Binary, 1 = Positive, 0 = Negative
c. **Verified**, *verified*: Binary, 1 = Yes, 0 = No

Cross-tabulations of these data sets are given in Figure 2.

**Fig 2.**
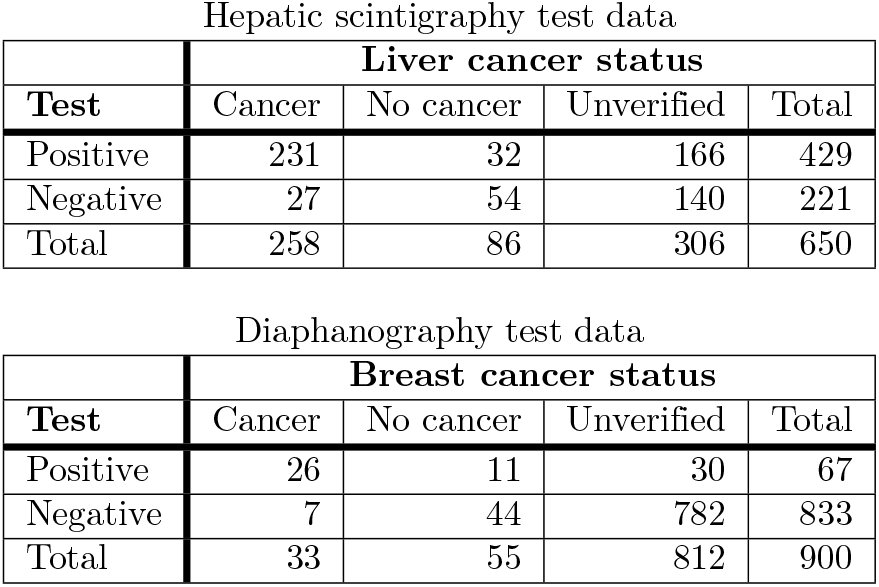
Cross-tabulation of hepatic scintigraphy and diaphanography data sets.

### Proposed methods

#### Scaled inverse probability weighted resampling

Based on the IPB method proposed by Arifin and Yusof [21], the scaled inverse probability weighted resampling (SIPW) method is proposed to overcome the shortcomings of IPB for correcting PVB. The proposed SIPW algorithm is given in Algorithm 1. IPB defines *n* as the observed sample size. In contrast, SIPW defines *n* as the full sample size that included both verified and unverified samples. The verified sample size is denoted as *n*_1_, while the unverified sample size is denoted as *n*_0_. A sample of size *n* is sampled with replacement for *b* times from *n*_1_, resulting in *b* samples. Because SIPW does not perform sampling with replacement to obtain the original *n*_1_ observed sample size, this is no longer a bootstrap sample as seen in IPB. Thus, the general term, *resampling*, is used in the name of the method.

#### Scaled inverse probability weighted balanced resampling

During the course of this research, it was observed that subgroup sample sizes, *n*_*D*=1_ and *n*_*D*=0_ for diseased and non-diseased observations respectively, directly affect the precision (i.e. SE) of Sn or Sp as a smaller sample size results in lower precision (higher SE). The SE, in particular for Sn, can increase even further when the disease prevalence gets lower as the subgroup size of *n*_*D*=1_ will also get smaller. In order to reduce the SE, a possible solution was inspired by how a diagnostic accuracy study is designed.

A diagnostic accuracy study is ideally designed as a cohort study [3, 4, 31]. Whenever the disease is very rare, a case-control study is more feasible, where those with the disease (case group) are compared to those without the disease (control group), often using a 1:1 to 1:3 ratio [31, 32]. This approach is similar to data-level resampling methods commonly used to address the class imbalance problem in machine learning, where the class distribution is rebalanced through over- or under-sampling methods [33].

Therefore, the scaled inverse probability weighted balanced resampling (SIPW-B) method is proposed to mimic the case-control study design. SIPW-B balances the effect of subgroup sample sizes by resizing the subgroup sizes to a predefined ratio of *n*_*D*=0_:*n*_*D*=1_, while keeping the original sample size *n* = *n*_*D*=0_ + *n*_*D*=1_. SIPW-B algorithm is given in Algorithm 2. In the algorithm, the target control:case ratio or *n*_*D*=0_ : *n*_*D*=1_ is achieved through the following steps:

1. Set the desired relative size or ratio of control:case.
2. Calculate the initial relative size of *n*_*D*=0_ to *n*_*D*=1_ in the PVB sample.
3. Update *IPW*_*i*_ for instances *D*_*i*_ = 1 by multiplying the value with the initial relative size.
4. Calculate *k* as the sum of *IPW*_*i*_ for instances *D*_*i*_ = 0, divided by sum of *IPW*_*i*_ for instances *D*_*i*_ = 1.
5. Update *IPW*_*i*_ for instances *D*_*i*_ = 1 by multiplying the value with 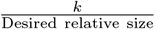
6. Calculate *SIPW*_*i*_ as *IPW*_*i*_ divided by the sum of *IPW*_*i*_.

For the experiments in this study, the target relative size of control:case was set at 1 or a ratio 1:1.

##### Algorithm 1 Scaled Inverse Probability Weighted Resampling

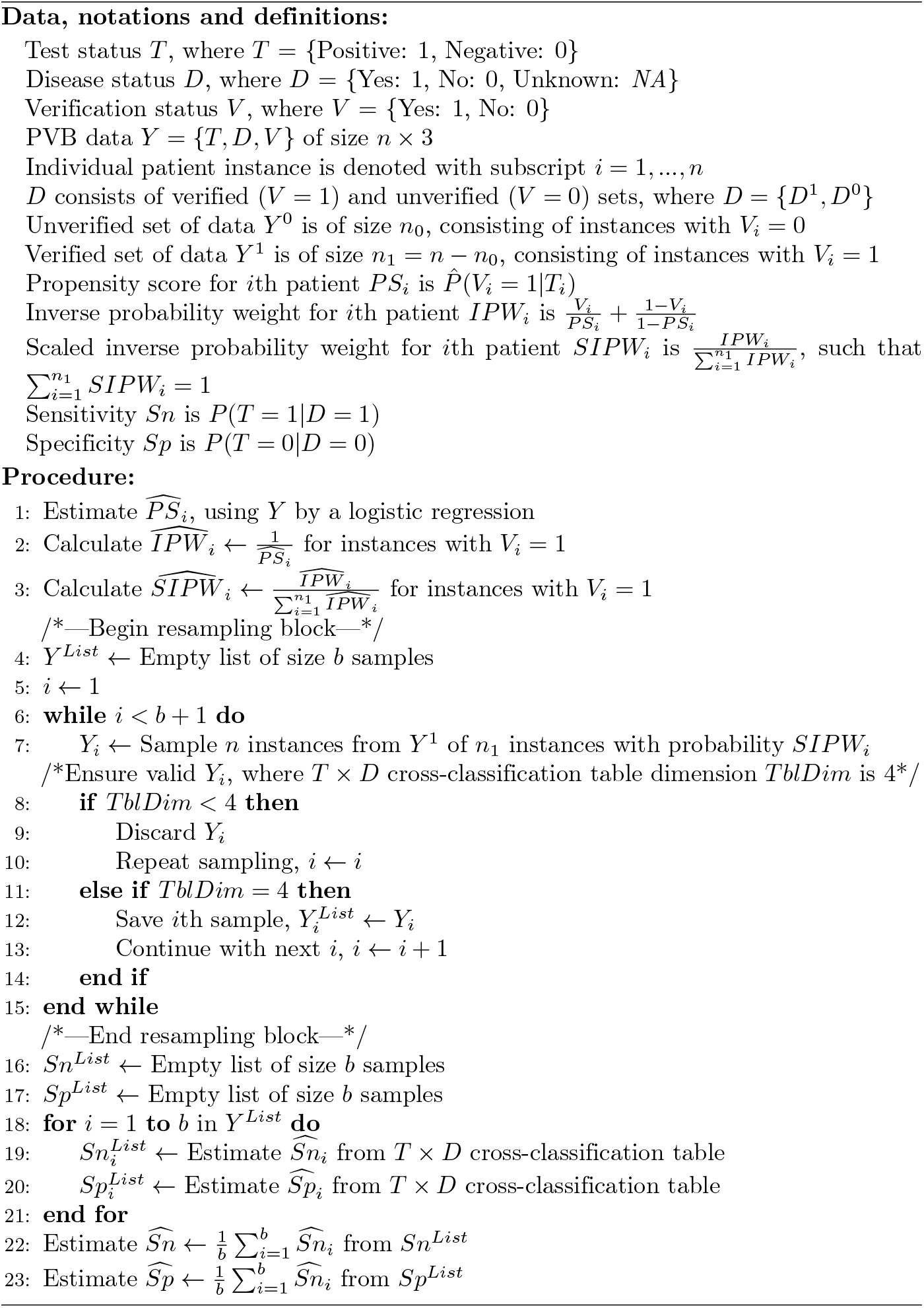

##### Algorithm 2 Scaled Inverse Probability Weighted Balanced Resampling

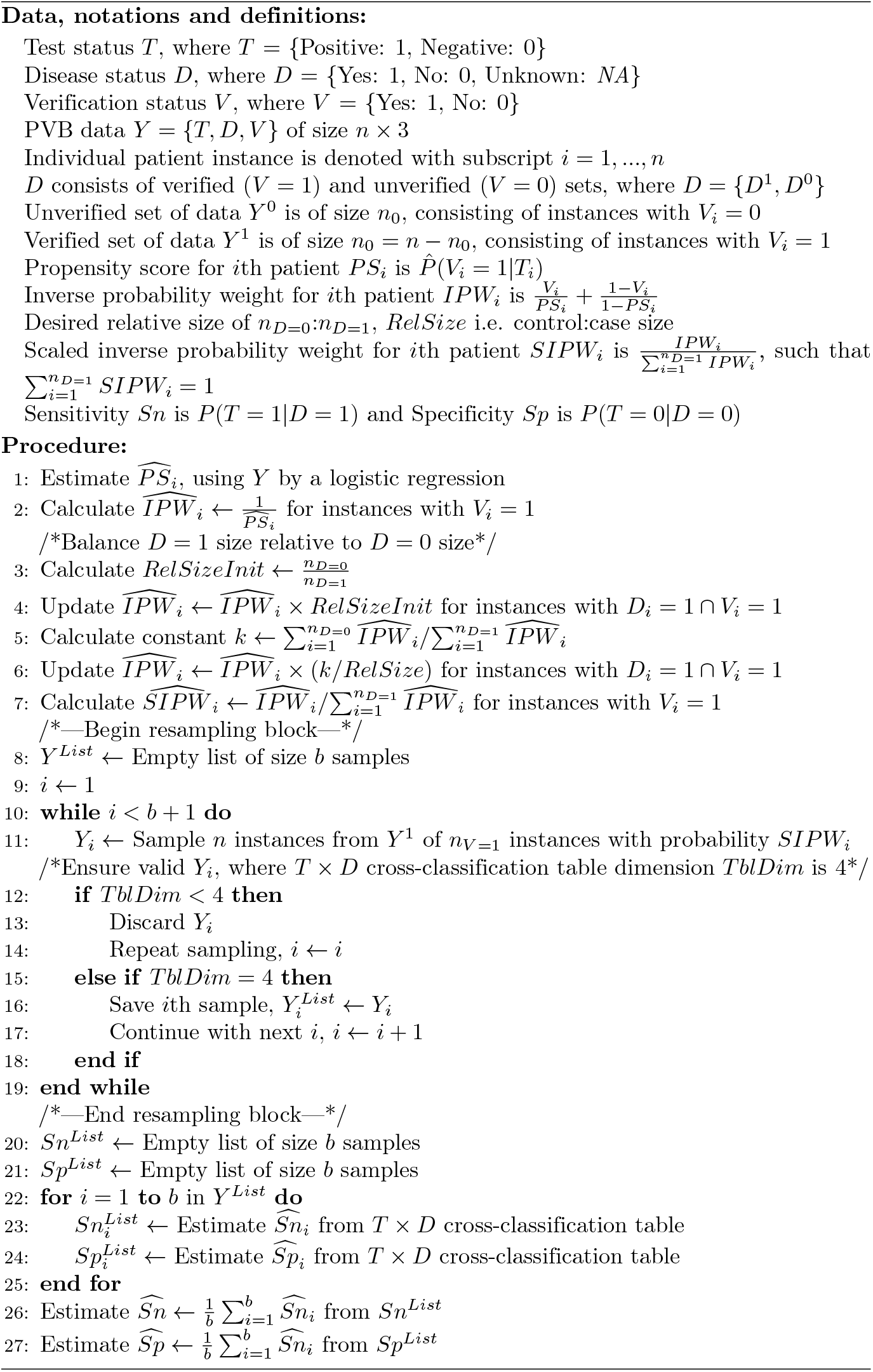

### Performance evaluation

#### Performance metrics

The performance evaluation relied on metrics that assess the difference between an estimate and its true value [22, 34, 35]. For a finite number of simulations *B*, the selected metrics are calculated as follows:

##### 1. Bias

Bias of a point estimator 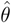 is defined as the difference between the expected value of 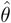 and the true value of a parameter *θ* [35]. *It is calculated as follows:*

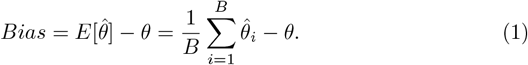

##### 2. Standard error

Standard error (SE) is the square root of the variance and is calculated as follows:

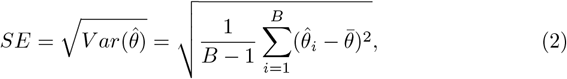

where 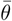 is the mean of 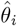 across repetitions.

Bias is often the primary metric of interest [22], as it indicates the accuracy of a method [35] and whether, on average, the method targets the parameter *θ* [22]. SE indicates the precision of the method [22, 35], with a smaller SE indicating higher precision [35].

#### Methods for comparison

The proposed SIPW and SIPW-B were compared with selected existing PVB correction methods, which are the BG method (representing the BG-based methods), the inverse probability weighting estimator (IPWE) and IPB methods (representing the PS-based methods) and the MI method (representing the imputation-based methods) were selected to represent different approaches for PVB correction. In addition, two additional methods were also compared; full data analysis (FDA) as the ideal method whenever full and unbiased data are available [3], and complete case analysis (CCA) as the uncorrected method that is biased in the presence of PVB [36]. The details of Sn and Sp calculation for these comparison methods were already described elsewhere [21, 37].

For the simulated datasets, the methods are compared based on the mean of the estimates, bias and SE, organized by sample sizes and Sn-Sp combinations. Coverage, defined as the proportion of times the confidence interval includes the true estimate [22, 34, 35], was not included as a performance metric for comparing the methods in our simulation. This was because, we did not propose a new method to obtain the CI for SIPW and SIPW-B.

For the clinical data sets, point estimates and the respective 95% CIs were compared. For CCA, the CIs for Sn and Sp were calculated by using Wald interval, while for the BG method, the calculation step given in the original article was followed [38]. For IPWE, IPB, SIPW and SIPW-B, the CIs were obtained by bootstrap percentile interval method [39]. For MI, the CIs were obtained by Rubin’s rule [37, 40].

#### Experimental setup

R statistical programming language [41] version 3.6.3 was used to run the experiments within RStudio [42] integrated development environment. The choice of the final stable version of the 3.x.x versions of the R language was to ensure reproducibility of the experimental results compared to the currently used 4.x.x versions of the R language which is still in continuous development. *mice* [43] version 3.14.0 and *simstudy* [44] version 0.5.0 R packages were used. The seed number for the random number generator was set to 3209673. For simulated data sets, the settings were the number of simulation runs *B* = 500, samples *b* = 1000 (for IPB, SIPW and SIPW-B) and imputations *m* = 100 [45, 46]. For clinical data sets, the settings were the number of samples *b* = 1000 (for IPB, SIPW and SIPW-B), bootstrap replicates *R* = 1000 (to obtain the CIs for IPWE, SIPW and SIPW-B) and *m* = the percentage of incomplete cases for real clinical data sets [47–49]. Please note that IPB does not need an additional round of bootstrapping to obtain its CI, as bootstrap is integral to the method itself.

## Results

### Simulated data sets

The simulation results for the FDA, CCA and PVB correction methods for *p* = 0.4 are displayed in Table 1, comparing sample sizes *n* = 200 and 1000. The results are arranged by Sn = (0.3, 0.6, 0.9) and Sp = (0.6, 0.9) parameter combinations. The proportions of verification *P*(*V* = 1) were 0.54, 0.47, 0.59, 0.52, 0.64 and 0.57 for (0.3, 0.6), (0.3, 0.9), (0.6, 0.6), (0.6, 0.9), (0.9, 0.6) and (0.9, 0.9) for (Sn, Sp) pairs respectively. The best values (i.e. the smallest bias and SE values) achieved by IPB, SIPW, and SIPW-B are marked with an asterisk.

**Table 1.** Comparison between IPB and existing PVB correction methods for *p* = 0.4 with *n* = 200 and 1000 under six combinations of Sn and Sp.

| Methods | Mean | Bias | SE | Mean | Bias | SE | Mean | Bias | SE | Mean | Bias | SE |
| --- | --- | --- | --- | --- | --- | --- | --- | --- | --- | --- | --- | --- |
| | $n = 200$ | | | | | | $n = 1000$ | | | | | |
|  | Sn = 0.3 |  |  | Sp = 0.6 |  |  | Sn = 0.3 |  |  | Sp = 0.6 |  |  |
| FDA | 0.302 | 0.002 | 0.050 | 0.603 | 0.003 | 0.044 | 0.302 | 0.002 | 0.023 | 0.600 | 0.000 | 0.019 |
| CCA | 0.465 | 0.165 | 0.076 | 0.430 | -0.170 | 0.060 | 0.465 | 0.165 | 0.036 | 0.429 | -0.171 | 0.027 |
| BG | 0.303 | 0.003 | 0.058 | 0.602 | 0.002 | 0.051 | 0.303 | 0.003 | 0.028 | 0.601 | 0.001 | 0.023 |
| IPWE | 0.303 | 0.003 | 0.058 | 0.602 | 0.002 | 0.051 | 0.303 | 0.003 | 0.028 | 0.601 | 0.001 | 0.023 |
| MI | 0.305 | 0.005 | 0.060 | 0.600 | 0.000 | 0.053 | 0.303 | 0.003 | 0.029 | 0.600 | 0.000 | 0.023 |
| IPB | 0.300 | *0.000 | 0.085 | 0.596 | *-0.004 | 0.080 | 0.302 | *0.002 | 0.042 | 0.600 | *0.000 | 0.035 |
| SIPW | 0.302 | 0.002 | 0.079 | 0.604 | *0.004 | 0.070 | 0.302 | *0.002 | 0.036 | 0.601 | 0.001 | *0.031 |
| SIPW-B | 0.302 | 0.002 | *0.078 | 0.605 | 0.005 | *0.067 | 0.304 | 0.004 | *0.035 | 0.602 | 0.002 | 0.032 |
|  | Sn = 0.3 |  |  | Sp = 0.9 |  |  | Sn = 0.3 |  |  | Sp = 0.9 |  |  |
| FDA | 0.302 | 0.002 | 0.050 | 0.902 | 0.002 | 0.027 | 0.302 | 0.002 | 0.023 | 0.900 | 0.000 | 0.013 |
| CCA | 0.466 | 0.166 | 0.077 | 0.821 | -0.079 | 0.053 | 0.464 | 0.164 | 0.034 | 0.818 | -0.082 | 0.024 |
| BG | 0.305 | 0.005 | 0.060 | 0.902 | 0.002 | 0.030 | 0.302 | 0.002 | 0.026 | 0.900 | 0.000 | 0.014 |
| IPWE | 0.305 | 0.005 | 0.060 | 0.902 | 0.002 | 0.030 | 0.302 | 0.002 | 0.026 | 0.900 | 0.000 | 0.014 |
| MI | 0.306 | 0.006 | 0.061 | 0.901 | 0.001 | 0.030 | 0.302 | 0.002 | 0.027 | 0.900 | 0.000 | 0.014 |
| IPB | 0.306 | *0.006 | 0.095 | 0.903 | 0.003 | 0.050 | 0.301 | *0.001 | 0.043 | 0.900 | *0.000 | 0.022 |
| SIPW | 0.307 | 0.007 | 0.081 | 0.902 | *0.002 | *0.041 | 0.302 | 0.002 | 0.035 | 0.901 | 0.001 | *0.018 |
| SIPW-B | 0.308 | 0.008 | *0.076 | 0.905 | 0.005 | 0.042 | 0.301 | *0.001 | *0.034 | 0.900 | *0.000 | 0.019 |
|  | Sn = 0.6 |  |  | Sp = 0.6 |  |  | Sn = 0.6 |  |  | Sp = 0.6 |  |  |
| FDA | 0.603 | 0.003 | 0.055 | 0.602 | 0.002 | 0.044 | 0.602 | 0.002 | 0.023 | 0.600 | 0.000 | 0.019 |
| CCA | 0.754 | 0.154 | 0.060 | 0.430 | -0.170 | 0.060 | 0.751 | 0.151 | 0.026 | 0.428 | -0.172 | 0.026 |
| BG | 0.607 | 0.007 | 0.072 | 0.602 | 0.002 | 0.050 | 0.602 | 0.002 | 0.031 | 0.600 | 0.000 | 0.022 |
| IPWE | 0.607 | 0.007 | 0.072 | 0.602 | 0.002 | 0.050 | 0.602 | 0.002 | 0.031 | 0.600 | 0.000 | 0.022 |
| MI | 0.605 | 0.005 | 0.075 | 0.599 | -0.001 | 0.052 | 0.601 | 0.001 | 0.032 | 0.599 | -0.001 | 0.022 |
| IPB | 0.609 | 0.009 | 0.105 | 0.602 | 0.002 | 0.078 | 0.599 | *-0.001 | 0.044 | 0.601 | 0.001 | 0.034 |
| SIPW | 0.608 | 0.008 | 0.090 | 0.603 | 0.003 | *0.065 | 0.604 | 0.004 | *0.037 | 0.600 | *0.000 | *0.029 |
| SIPW-B | 0.606 | *0.006 | *0.084 | 0.600 | *0.000 | 0.070 | 0.602 | 0.002 | 0.038 | 0.600 | *0.000 | 0.031 |
|  | Sn = 0.6 |  |  | Sp = 0.9 |  |  | Sn = 0.6 |  |  | Sp = 0.9 |  |  |
| FDA | 0.603 | 0.003 | 0.055 | 0.902 | 0.002 | 0.027 | 0.602 | 0.002 | 0.023 | 0.900 | 0.000 | 0.012 |
| CCA | 0.754 | 0.154 | 0.061 | 0.822 | -0.078 | 0.054 | 0.752 | 0.152 | 0.026 | 0.818 | -0.082 | 0.025 |
| BG | 0.608 | 0.008 | 0.075 | 0.903 | 0.003 | 0.030 | 0.602 | 0.002 | 0.031 | 0.900 | 0.000 | 0.014 |
| IPWE | 0.608 | 0.008 | 0.075 | 0.903 | 0.003 | 0.030 | 0.602 | 0.002 | 0.031 | 0.900 | 0.000 | 0.014 |
| MI | 0.605 | 0.005 | 0.076 | 0.901 | 0.001 | 0.031 | 0.602 | 0.002 | 0.032 | 0.900 | 0.000 | 0.014 |
| IPB | 0.605 | *0.005 | 0.118 | 0.903 | *0.003 | 0.049 | 0.599 | *-0.001 | 0.048 | 0.899 | -0.001 | 0.024 |
| SIPW | 0.610 | 0.010 | 0.094 | 0.903 | *0.003 | *0.040 | 0.604 | 0.004 | 0.040 | 0.901 | 0.001 | *0.019 |
| SIPW-B | 0.608 | 0.008 | *0.086 | 0.905 | 0.005 | 0.042 | 0.602 | 0.002 | *0.039 | 0.900 | *0.000 | 0.020 |
|  | Sn = 0.9 |  |  | Sp = 0.6 |  |  | Sn = 0.9 |  |  | Sp = 0.6 |  |  |
| FDA | 0.899 | -0.001 | 0.033 | 0.601 | 0.001 | 0.044 | 0.901 | 0.001 | 0.015 | 0.601 | 0.001 | 0.020 |
| CCA | 0.945 | 0.045 | 0.027 | 0.427 | -0.173 | 0.057 | 0.948 | 0.048 | 0.012 | 0.429 | -0.171 | 0.027 |
| BG | 0.896 | -0.004 | 0.046 | 0.600 | 0.000 | 0.046 | 0.901 | 0.001 | 0.022 | 0.601 | 0.001 | 0.021 |
| IPWE | 0.896 | -0.004 | 0.046 | 0.600 | 0.000 | 0.046 | 0.901 | 0.001 | 0.022 | 0.601 | 0.001 | 0.021 |
| MI | 0.889 | -0.011 | 0.046 | 0.598 | -0.002 | 0.047 | 0.899 | -0.001 | 0.023 | 0.600 | 0.000 | 0.021 |
| IPB | 0.894 | -0.006 | 0.064 | 0.595 | -0.005 | 0.072 | 0.901 | *0.001 | 0.028 | 0.600 | *0.000 | 0.033 |
| SIPW | 0.896 | -0.004 | 0.058 | 0.597 | -0.003 | *0.064 | 0.901 | *0.001 | 0.027 | 0.600 | *0.000 | *0.029 |
| SIPW-B | 0.897 | *-0.003 | *0.055 | 0.599 | *-0.001 | 0.069 | 0.901 | *0.001 | *0.025 | 0.603 | 0.003 | 0.031 |
|  | Sn = 0.9 |  |  | Sp = 0.9 |  |  | Sn = 0.9 |  |  | Sp = 0.9 |  |  |
| FDA | 0.899 | -0.001 | 0.033 | 0.900 | 0.000 | 0.028 | 0.901 | 0.001 | 0.015 | 0.901 | 0.001 | 0.012 |
| CCA | 0.946 | 0.046 | 0.028 | 0.817 | -0.083 | 0.054 | 0.948 | 0.048 | 0.013 | 0.819 | -0.081 | 0.024 |
| BG | 0.899 | -0.001 | 0.048 | 0.900 | 0.000 | 0.030 | 0.901 | 0.001 | 0.022 | 0.901 | 0.001 | 0.013 |
| IPWE | 0.899 | -0.001 | 0.048 | 0.900 | 0.000 | 0.030 | 0.901 | 0.001 | 0.022 | 0.901 | 0.001 | 0.013 |
| MI | 0.889 | -0.011 | 0.050 | 0.898 | -0.002 | 0.031 | 0.899 | -0.001 | 0.023 | 0.901 | 0.001 | 0.013 |
| IPB | 0.899 | -0.001 | 0.066 | 0.901 | 0.001 | 0.045 | 0.901 | *0.001 | 0.031 | 0.901 | *0.001 | 0.020 |
| SIPW | 0.900 | *0.000 | 0.060 | 0.900 | *0.000 | *0.041 | 0.902 | 0.002 | *0.026 | 0.901 | *0.001 | *0.019 |
| SIPW-B | 0.898 | -0.002 | *0.057 | 0.898 | -0.002 | 0.044 | 0.902 | 0.002 | *0.026 | 0.901 | *0.001 | *0.019 |

Abbreviations: BG, Begg and Greenes' method; CCA, complete case analysis; FDA, Full data analysis; IPB, inverse probability bootstrap; IPWE, inverse probability weighting estimator; MI, multiple imputation; $n$ , sample size; $p$ , disease prevalence; SE, standard error; SIPW, scaled inverse probability weighted resampling; SIPW-B, scaled inverse probability weighted balanced resampling; Sn, sensitivity; Sp, specificity.

Next, The simulation results for the FDA, CCA and PVB correction methods for *p* = 0.1 are displayed in Table 2 for sample sizes *n* = 200 and 1000. The results are arranged by Sn = (0.3, 0.6, 0.9) and Sp = (0.6, 0.9) parameter combinations. The proportions of verification *P*(*V* = 1) were 0.56, 0.45, 0.57, 0.46, 0.58 and 0.472 for (0.3, 0.6), (0.3, 0.9), (0.6, 0.6), (0.6, 0.9), (0.9, 0.6) and (0.9, 0.9) for (Sn, Sp) pairs respectively. Again, the smallest bias and SE values achieved by IPB, SIPW, and SIPW-B are marked with an asterisk.

**Table 2.** Comparison between IPB and existing PVB correction methods for *p* = 0.1 with *n* = 200 and 1000 under six combinations of Sn and Sp.

| Methods | Mean | Bias | SE | Mean | Bias | SE | Mean | Bias | SE | Mean | Bias | SE |
| --- | --- | --- | --- | --- | --- | --- | --- | --- | --- | --- | --- | --- |
| | $n = 200$ | | | | | | $n = 1000$ | | | | | |
|  | Sn = 0.3 |  |  | Sp = 0.6 |  |  | Sn = 0.3 |  |  | Sp = 0.6 |  |  |
| FDA | 0.301 | 0.001 | 0.107 | 0.601 | 0.001 | 0.037 | 0.300 | 0.000 | 0.043 | 0.600 | 0.000 | 0.016 |
| CCA | 0.459 | 0.159 | 0.159 | 0.428 | -0.172 | 0.050 | 0.462 | 0.162 | 0.066 | 0.429 | -0.171 | 0.022 |
| BG | 0.310 | 0.010 | 0.141 | 0.600 | 0.000 | 0.039 | 0.302 | 0.002 | 0.055 | 0.600 | 0.000 | 0.017 |
| IPWE | 0.310 | 0.010 | 0.141 | 0.600 | 0.000 | 0.039 | 0.302 | 0.002 | 0.055 | 0.600 | 0.000 | 0.017 |
| MI | 0.303 | 0.003 | 0.133 | 0.598 | -0.002 | 0.039 | 0.303 | 0.003 | 0.055 | 0.600 | 0.000 | 0.017 |
| IPB | 0.309 | 0.009 | 0.209 | 0.601 | *0.001 | 0.063 | 0.303 | 0.003 | 0.080 | 0.600 | *0.000 | 0.027 |
| SIPW | 0.306 | *0.006 | 0.167 | 0.599 | *-0.001 | *0.052 | 0.308 | 0.008 | 0.071 | 0.599 | -0.001 | *0.024 |
| SIPW-B | 0.307 | 0.007 | *0.148 | 0.601 | *0.001 | 0.060 | 0.300 | *0.000 | *0.058 | 0.599 | -0.001 | 0.028 |
|  | Sn = 0.3 |  |  | Sp = 0.9 |  |  | Sn = 0.3 |  |  | Sp = 0.9 |  |  |
| FDA | 0.298 | -0.002 | 0.106 | 0.901 | 0.001 | 0.021 | 0.300 | 0.000 | 0.043 | 0.900 | 0.000 | 0.010 |
| CCA | 0.464 | 0.164 | 0.160 | 0.819 | -0.081 | 0.042 | 0.461 | 0.161 | 0.068 | 0.818 | -0.082 | 0.018 |
| BG | 0.314 | 0.014 | 0.136 | 0.901 | 0.001 | 0.022 | 0.302 | 0.002 | 0.056 | 0.900 | 0.000 | 0.010 |
| IPWE | 0.314 | 0.014 | 0.136 | 0.901 | 0.001 | 0.022 | 0.302 | 0.002 | 0.056 | 0.900 | 0.000 | 0.010 |
| MI | 0.302 | 0.002 | 0.129 | 0.901 | 0.001 | 0.023 | 0.299 | -0.001 | 0.059 | 0.900 | 0.000 | 0.010 |
| IPB | 0.323 | 0.023 | 0.227 | 0.903 | 0.003 | 0.039 | 0.305 | 0.005 | 0.092 | 0.900 | *0.000 | 0.019 |
| SIPW | 0.310 | *0.010 | 0.178 | 0.901 | *0.001 | *0.033 | 0.304 | 0.004 | 0.074 | 0.900 | *0.000 | *0.014 |
| SIPW-B | 0.318 | 0.018 | *0.143 | 0.904 | 0.004 | 0.037 | 0.302 | *0.002 | *0.061 | 0.900 | *0.000 | 0.017 |
|  | Sn = 0.6 |  |  | Sp = 0.6 |  |  | Sn = 0.6 |  |  | Sp = 0.6 |  |  |
| FDA | 0.596 | -0.004 | 0.112 | 0.601 | 0.001 | 0.037 | 0.600 | 0.000 | 0.048 | 0.600 | 0.000 | 0.017 |
| CCA | 0.743 | 0.143 | 0.117 | 0.429 | -0.171 | 0.049 | 0.754 | 0.154 | 0.053 | 0.429 | -0.171 | 0.022 |
| BG | 0.603 | 0.003 | 0.144 | 0.601 | 0.001 | 0.038 | 0.607 | 0.007 | 0.067 | 0.601 | 0.001 | 0.017 |
| IPWE | 0.603 | 0.003 | 0.144 | 0.601 | 0.001 | 0.038 | 0.607 | 0.007 | 0.067 | 0.601 | 0.001 | 0.017 |
| MI | 0.579 | -0.021 | 0.136 | 0.598 | -0.002 | 0.039 | 0.601 | 0.001 | 0.067 | 0.600 | 0.000 | 0.017 |
| IPB | 0.595 | -0.005 | 0.202 | 0.606 | 0.006 | 0.062 | 0.605 | 0.005 | 0.093 | 0.600 | *0.000 | 0.027 |
| SIPW | 0.613 | 0.013 | 0.184 | 0.603 | 0.003 | *0.055 | 0.604 | *0.004 | 0.084 | 0.600 | *0.000 | *0.025 |
| SIPW-B | 0.600 | *0.000 | *0.157 | 0.601 | *0.001 | 0.062 | 0.607 | 0.007 | *0.071 | 0.601 | 0.001 | 0.027 |
|  | Sn = 0.6 |  |  | Sp = 0.9 |  |  | Sn = 0.6 |  |  | Sp = 0.9 |  |  |
| FDA | 0.600 | 0.000 | 0.115 | 0.900 | 0.000 | 0.022 | 0.600 | 0.000 | 0.048 | 0.900 | 0.000 | 0.010 |
| CCA | 0.738 | 0.138 | 0.119 | 0.818 | -0.082 | 0.043 | 0.749 | 0.149 | 0.052 | 0.819 | -0.081 | 0.019 |
| BG | 0.598 | -0.002 | 0.143 | 0.900 | 0.000 | 0.023 | 0.601 | 0.001 | 0.065 | 0.900 | 0.000 | 0.010 |
| IPWE | 0.598 | -0.002 | 0.143 | 0.900 | 0.000 | 0.023 | 0.601 | 0.001 | 0.065 | 0.900 | 0.000 | 0.010 |
| MI | 0.568 | -0.032 | 0.134 | 0.900 | 0.000 | 0.023 | 0.593 | -0.007 | 0.065 | 0.900 | 0.000 | 0.010 |
| IPB | 0.599 | -0.001 | 0.214 | 0.898 | -0.002 | 0.042 | 0.604 | 0.004 | 0.099 | 0.901 | 0.001 | 0.018 |
| SIPW | 0.600 | *0.000 | 0.178 | 0.899 | *-0.001 | *0.033 | 0.602 | 0.002 | 0.080 | 0.901 | 0.001 | *0.014 |
| SIPW-B | 0.598 | -0.002 | *0.152 | 0.902 | 0.002 | 0.038 | 0.600 | *0.000 | *0.067 | 0.900 | *0.000 | 0.017 |
|  | Sn = 0.9 |  |  | Sp = 0.6 |  |  | Sn = 0.9 |  |  | Sp = 0.6 |  |  |
| FDA | 0.875 | -0.025 | 0.062 | 0.602 | 0.002 | 0.035 | 0.899 | -0.001 | 0.028 | 0.600 | 0.000 | 0.016 |
| CCA | 0.910 | 0.010 | 0.042 | 0.430 | -0.170 | 0.048 | 0.947 | 0.047 | 0.025 | 0.428 | -0.172 | 0.021 |
| BG | 0.837 | -0.063 | 0.068 | 0.599 | -0.001 | 0.037 | 0.900 | 0.000 | 0.044 | 0.600 | 0.000 | 0.017 |
| IPWE | 0.837 | -0.063 | 0.068 | 0.599 | -0.001 | 0.037 | 0.900 | 0.000 | 0.044 | 0.600 | 0.000 | 0.017 |
| MI | 0.800 | -0.100 | 0.080 | 0.597 | -0.003 | 0.037 | 0.889 | -0.011 | 0.046 | 0.600 | 0.000 | 0.017 |
| IPB | 0.832 | -0.068 | 0.133 | 0.605 | 0.005 | 0.059 | 0.897 | -0.003 | 0.060 | 0.601 | 0.001 | 0.027 |
| SIPW | 0.837 | *-0.063 | 0.108 | 0.600 | *0.000 | *0.050 | 0.901 | 0.001 | 0.052 | 0.600 | *0.000 | *0.023 |
| SIPW-B | 0.837 | *-0.063 | *0.077 | 0.600 | *0.000 | 0.061 | 0.900 | *0.000 | *0.045 | 0.601 | 0.001 | 0.028 |
|  | Sn = 0.9 |  |  | Sp = 0.9 |  |  | Sn = 0.9 |  |  | Sp = 0.9 |  |  |
| FDA | 0.874 | -0.026 | 0.062 | 0.901 | 0.001 | 0.023 | 0.900 | 0.000 | 0.029 | 0.900 | 0.000 | 0.010 |
| CCA | 0.905 | 0.005 | 0.051 | 0.819 | -0.081 | 0.045 | 0.948 | 0.048 | 0.025 | 0.819 | -0.081 | 0.020 |
| BG | 0.830 | -0.070 | 0.079 | 0.900 | 0.000 | 0.025 | 0.901 | 0.001 | 0.044 | 0.900 | 0.000 | 0.011 |
| IPWE | 0.830 | -0.070 | 0.079 | 0.900 | 0.000 | 0.025 | 0.901 | 0.001 | 0.044 | 0.900 | 0.000 | 0.011 |
| MI | 0.777 | -0.123 | 0.087 | 0.899 | -0.001 | 0.025 | 0.887 | -0.013 | 0.045 | 0.900 | 0.000 | 0.011 |
| IPB | 0.829 | -0.071 | 0.158 | 0.900 | *0.000 | 0.044 | 0.898 | -0.002 | 0.061 | 0.900 | *0.000 | 0.018 |
| SIPW | 0.831 | -0.069 | 0.112 | 0.900 | *0.000 | *0.034 | 0.901 | *0.001 | 0.055 | 0.900 | *0.000 | *0.015 |
| SIPW-B | 0.832 | *-0.068 | *0.089 | 0.900 | *0.000 | 0.037 | 0.901 | *0.001 | *0.046 | 0.900 | *0.000 | 0.016 |

Abbreviations: BG, Begg and Greenes' method; CCA, complete case analysis; FDA, Full data analysis; IPB, inverse probability bootstrap; IPWE, inverse probability weighting estimator; MI, multiple imputation; $n$ , sample size; $p$ , disease prevalence; SE, standard error; SIPW, scaled inverse probability weighted resampling; SIPW-B, scaled inverse probability weighted balanced resampling; Sn, sensitivity; Sp, specificity.

As observed in Table 1 and 2, both SIPW and SIPW-B generally performed better than IPB for Sn and Sp estimation as indicated by smaller bias and SE values. SIPW-B performed better (smaller SE) for Sn compared to SIPW by enlarging the size of the case group (*n*_*D*=1_). As expected, SIPW showed smaller SE than SIPW-B for Sp estimation because it maintains the original size of the control group (*n*_*D*=0_), of which at a disease prevalence of *p* = 0.4, the control group size is larger than the case group (Table 1). This effect was more notable at a lower disease prevalence of *p* = 0.1 (Table 2). Bias values showed mixed results, with marginal differences observed among IPB, SIPW, and SIPW-B. When compared to the existing methods (BG, IPWE and MI), SIPW-B closely matched these methods for the Sn, while SIPW closely aligned with the results for these methods for Sp estimation. A further observation was made that across all PVB correction methods, both bias and SE decreased with increasing disease prevalence and sample size. As a side note, when the prevalence is low and the sample size is small, CCA turned out to be less biased than FDA and other PVB correction methods at a very high Sn value of 0.9, which was unexpected.

### Clinical data sets

The results comparing CCA (bias uncorrected) and the PVB correction methods using the clinical data sets are displayed in Table 3. All PVB correction methods showed almost identical results for Sn and Sp point estimates across these data sets, with the exception of MI, which showed a slightly different Sn estimate for the diaphanography data set. In contrast to IPB, for the hepatic data set, SIPW and SIPW-B showed 95% CIs that were more consistent with existing methods. The diaphanography data set further highlighted the limitations of IPB, as its 95% CIs for Sn and Sp were notably wider and diverged from those estimated by SIPW, SIPW-B and existing methods. Based on the small observed sample size of this data set (only 88 verified out of 900 patients), IPB appears to perform poorly with interval estimation in situations with limited data.

**Table 3.** Sn and Sp estimates of IPB, SIPW, SIPW-B and other methods with the respective 95% CIs using clinical data sets.

| Methods | Scintigraphy data set |  | Diaphanography data set |  |
| --- | --- | --- | --- | --- |
|  | Sn (95% CI) | Sp (95% CI) | Sn (95% CI) | Sp (95% CI) |
| CCA | 0.895 (0.858, 0.933) | 0.628 (0.526, 0.730) | 0.788 (0.648, 0.927) | 0.800 (0.694, 0.906) |
| BG | 0.836 (0.788, 0.884) | 0.738 (0.662, 0.815) | 0.292 (0.134, 0.449) | 0.973 (0.958, 0.988) |
| IPWE | 0.836 (0.785, 0.885) | 0.738 (0.656, 0.812) | 0.292 (0.177, 0.548) | 0.973 (0.957, 0.987) |
| MI | 0.834 (0.782, 0.885) | 0.738 (0.661, 0.815) | 0.279 (0.124, 0.435) | 0.972 (0.957, 0.987) |
| IPB | 0.838 (0.793, 0.881) | 0.738 (0.650, 0.824) | 0.290 (0.077, 0.529) | 0.973 (0.931, 1.000) |
| SIPW | 0.837 (0.785, 0.886) | 0.739 (0.655, 0.811) | 0.292 (0.176, 0.548) | 0.973 (0.957, 0.987) |
| SIPW-B | 0.837 (0.785, 0.885) | 0.739 (0.655, 0.812) | 0.291 (0.176, 0.548) | 0.973 (0.957, 0.987) |

Abbreviations: BG, Begg and Greenes’ method; CCA, complete case analysis; CI, confidence interval; IPB, inverse probability bootstrap; IPWE, inverse probability weighting estimator; MI, multiple imputation; SIPW, scaled inverse probability weighted resampling; SIPW-B, scaled inverse probability weighted balanced resampling; Sn, sensitivity; Sp, specificity.

## Discussion

This study introduced two new methods based on the IPB method: SIPW and SIPW-B. These were compared against IPB and other existing methods across simulated data sets and clinical data sets to evaluate their performance in estimating Sn and Sp. In simulated data sets, both SIPW and SIPW-B demonstrated smaller bias and standard error values than IPB for Sn and Sp estimation, respectively. Specifically, SIPW-B and SIPW outperformed IPB in Sn and Sp estimation, respectively. In clinical data sets, all methods generally showed consistent results with each other, except for the Diaphanography data set where IPB and MI diverged from others. For practical applications, when disease prevalence is low, SIPW is preferable for estimating Sp, whereas SIPW-B is better suited for Sn estimation.

There are several advantages of the newly proposed methods as compared to IPB, while sharing the other advantages of IPB as discussed in another study [21]. First, SIPW and SIPW-B share the same advantage with IPB and MI as they allow the use of any full data analytical methods. In addition, SIPW and SIPW-B aligns better with MI by restoring full data *N*, while IPB restores the data containing the complete cases *n* only. The ability to utilize full data approach is advantageous as shown in [50] for the kappa coefficient in diagnostic accuracy studies. Second, both proposed methods showed lower SEs as compared to IPB. As highlighted in [21], a major disadvantage of IPB is its large SE compared to other existing methods. SIPW and SIPW-B overcome this issue by having comparable SEs to these existing methods. Third, the proposed methods were similar to IPB in that they are less biased than MI at low disease prevalence and comparable to BG and IPWE in terms of bias [21], while showing smaller SEs than IPB. Fourth, these two methods are easy to implement compared to MI, where they require the estimation of PS values, followed by the weighted resampling procedure. In contrast, in implementing MI for the PVB correction, one must choose suitable imputations methods as the performance depends on the selected imputation method [40, 51]. Finally, for PS-based methods (IPWE, IPB, SIPW and SIPW-B), they share the same advantage of using PS; the probability of verification given test result, *P*(*V* = 1| *T* = *t*). In contrast, BG-based and MI methods depend on the accurate estimation of the probability of disease status given test result, *P*(*D* = *d* |*T* = *t*). PS-based methods, however, utilize the accurate estimation of *P*(*V* = 1| *T* = *t*) to perform the correction. This makes PS-based methods particularly advantageous when a case-control study design is employed for diagnostic accuracy studies, as *P*(*D* = *d* | *T* = *t*) can be inaccurate in this situation [8].

However, there are several disadvantages of the proposed methods that can be highlighted. SIPW and SIPW-B are computationally intensive methods that rely on repeated resampling. Unlike IPB, CIs cannot be derived directly from these resamples because they no longer constitute valid bootstrap samples. For interval estimation, an additional computationally demanding bootstrapping procedure must be performed on top of the algorithms. Specifically, if 1000 bootstrap replicates (*R* = 1000) are required for interval estimation, the computational burden significantly increases. For instance, if 1000 bootstrap replicates (*R* = 1000) are needed for interval estimation, the time taken to complete a full SIPW procedure (e.g., 10 seconds with *b* = 1000 SIPW samples) will be multiplied by 1000. In addition, SIPW-B was designed to mimic the case-control study design. SIPW-B balances the effect of subgroup sample sizes by resizing the subgroup sizes to a predefined ratio of *n*_*D*=0_:*n*_*D*=1_, while keeping the original sample size *n* = *n*_*D*=0_ + *n*_*D*=1_. The side effect of the procedure is that it will lower the precision of the larger-sized subgroup, although the precision for the smaller-sized subgroup will increase. As an example, for a sample size *n* = 1000 with *p* = 0.2, the subgroup sizes will be *n*_*D*=1_ = *n× p* = 200 and *n*_*D*=0_ = *n×* (1 *- p*) = 800. The desired ratio of control:case is 1:1 ratio. Before the balancing procedure, Sn will be calculated from *n*_*D*=1_ = 200 while Sp from *n*_*D*=0_ = 800 sample sizes. After the balancing procedure, Sn will be calculated from *n*_*D*=1_ = 500 while Sp from *n*_*D*=0_ = 500 sample sizes. Notably, the subgroup size *n*_*D*=0_ reduces from 800 to 500 for the calculation of Sp, reducing both the counts for numerator and denominator, thus lowering the precision of Sp. On the positive note, the subgroup size *n*_*D*=1_ increases from 200 to 500 for the calculation of Sn, increasing both the counts for numerator and denominator, thus increasing the precision of Sn. Therefore, it is recommended to use SIPW-B only when Sn and Sp are the main estimates to be calculated.

Therefore, we recommend using SIPW when complete data restoration is needed for further analysis involving additional metrics than depends on prevalence like PPV, NPV and accuracy [3, 52], rather than just Sn and Sp. Conversely, SIPW-B should be employed only when the primary aim is to obtain accurate Sn and Sp estimates. After applying SIPW-B, which mimics a case-control study design, statistics that rely on an accurate distribution of *P*(*D* = 1) (such as PPV = *P*(*D* = 1| *T* = 1) or NPV = *P*(*D* = 0 |*T* = 0)) are no longer valid [8]. Although PPV and NPV can be calculated indirectly from Sn and Sp point estimates of SIPW-B by utilizing Bayes’ theorem [3, 37], further research must be done to investigate this issue.

## Conclusion

This paper proposes the SIPW and SIPW-B methods to overcome the limitations of the IPB method in the context of PVB correction under the MAR assumption for binary diagnostic tests. The results show that both SIPW and SIPW-B outperformed IPB and were consistent with existing PVB correction methods. Specifically, SIPW excelled in estimating Sp, while SIPW-B excelled in Sn estimation, particularly when disease prevalence is low. The proposed methods offer advantages such as complete data restoration for subsequent analysis and the use of PS that are not influenced by disease prevalence. Although the new methods currently require more computational resources, this should become less of an issue with advancements in computational power.

## Data Availability

The data and code can be found in https://github.com/wnarifin/sipw_in_pvb For simulated data, data generation method is described for replication of the simulation.

## Author contributions

Conceptualization, W.N.A. and U.K.Y.; methodology, W.N.A. and U.K.Y.; software, W.N.A.; validation, W.N.A. and U.K.Y.; formal analysis, W.N.A.; investigation, W.N.A.; resources, U.K.Y.; data curation, W.N.A.; writing—original draft preparation, W.N.A.; writing—review and editing, W.N.A. and U.K.Y.; supervision, U.K.Y.; project administration, W.N.A.; funding acquisition, W.N.A. and U.K.Y. All authors have read and agreed to the published version of the manuscript.

## Acknowledgments

We thank our colleagues at the School of Computer Sciences and the School of Medical Sciences, Universiti Sains Malaysia for their comments on the early findings of this study and this article’s draft.

## Supporting Information

The data and code presented in this article are available from this GitHub repository: https://github.com/wnarifin/sipw_in_pvb.

